# Diffuse generalized phenotype of hypertrophic cardiomyopathy: genetic spectrum and surgical correction

**DOI:** 10.64898/2026.08.06.26359755

**Authors:** Sergey Dzemeshkevich, Balashova Maria, Margarita Polyak, Svetlana Solovyeva, Elena Mershina, Natalia Kotlukova, Elena Zaklyazminskaya

## Abstract

**Introduction:** Hypertrophic cardiomyopathy (HCM) is characterized by clinical and genetic heterogeneity. Age of manifestation, clinical and anatomical phenotypes of HCM vary significantly. This study discusses genetic causes and reconstructive surgery results for patients with particular intracardiac phenotype - diffused generalized HCM (DG-HCM).

**Methods:** personal and familial medical history, general examination, 12-lead resting ECG, 24-hour ECG Holter monitoring, transthoracic and transesophageal EchoCG, cardiac MRI with gadolinium enhancement. A ten-gene panel was sequenced by IonTorrent PGM. Mutational screening in patients with suspected multisystemic diseases was performed by Sanger sequencing.

**Results:** 170 patients with obstructive HCM (oHCM) requesting genetic counseling and surgical correction of HCM were evaluated. We distinguished particular DG-HCM subtype of oHCM (diffuse hypertrophy of IVS, LV free walls, papillary muscles displaced towards the LV apex) in 34 patients; 31 out of 34 underwent open heart reconstructive surgery. Patients with DG-HCM were younger at the time of surgery, had higher risk of SCD, and connective tissue dysplasia of the mitral valve. Hemodynamics normalization was observed in 1, 3, and 5 years after surgery. Eighteen ICDs were implanted; five patients experienced appropriate shocks. The genetic spectrum was enriched up to 30% by multisystem disorders. Mutations in “sarcomeric” genes were detected in 15%.

**Conclusion:** Intracardiac phenotype of HCM may correlate with genetic cause and long-term prognosis. DG-HCM phenotype accounts for 20% oHCM patients and indications for open-heart surgery. Extended myectomy with parietal resection of papillary muscles and correction of mitral valve insufficiency provides long-term benefits for DG-HCM patients. Multisystem disorders in patients with DG-HCM should be of special attention.

Study was supported by research project FURG-2024-0004.

## Introduction

Detection of idiopathic wall thickening in the most of cases leads to the diagnosis of hypertrophic cardiomyopathy (HCM), which is the most common inherited disease with a prevalence of 1:200 in the general population [1]. HCM is caused by mutations in the genes encoding different components of the sarcomere. Many of the mutation carriers do not have a pronounced clinical phenotype, and their quality of life and life expectancy is not significantly affected [1, 2]. There are, however, a considerable number of carriers who suffer from progressive heart failure (HF) as a result of the prominent cardiac hypertrophy, hemodynamic changes, various arrhythmias, and risk of SCD. These individuals often require continuous conservative or surgical treatment. The latter usually consists of ICD implantation, septal myectomy, alcohol ablation, and orthotopic heart transplantation [3].

The number of hospitals with expertise in the surgical treatment of HCM is growing steadily, leading to the rapid accumulation of data on this subject and some new essential insights into the clinical appearance and pathogenesis of the disease [4]. As many as 65% of primary HCM cases are associated with mutations in the genes encoding sarcomeric proteins [5]. The clinical and genetic polymorphism of HCM is gaining increasing awareness. As a result, the treatment strategies for different variants (or subtypes) of cardiac hypertrophy are being developed.

Not only the clinical course but also the anatomic profile of cardiac hypertrophy is highly polymorphic [5]. There is no generally accepted classification of anatomic variants of cardiac hypertrophy in HCM. The basal obstructive form of HCM is well-defined and the most frequently encountered variant [6, 7]. It can be treated safely and effectively with extended myectomy in experienced centers, including even the low-symptomatic latent forms [6]. There are other anatomic phenotypes of HCM, and the number of publications on different anatomic variant is increasing [7, 8]. However, many aspects, such as the underlying genetic contribution, the natural course of the disease, and the efficiency of surgical techniques, have yet to be elucidated.

This observational study presents the results of the clinical and genetic evaluation of patients with a particular variant of LV obstruction, referred to as diffuse generalized HCM (DG-HCM). An advanced reconstructive surgical technique was implemented, allowing for the normalization of the intraventricular pressure gradient and to increase the diastolic volume of the LV.

## Materials and methods

Inclusion criteria. We evaluated 170 patients with entering diagnosis of hypertrophic cardiomyopathy (cardiac hypertrophy > 13 mm), with significant obstruction in the left ventricular outflow tract (LVOT) (LVOT PGr ≥ 50 mm).

We found a specific anatomic phenotype characterized by diffuse hypertrophy of the left ventricular (LV) free walls, interventricular septum, and papillary muscles, with their displacement towards the LV apex. The presence of these anatomic features was used to pick out and to characterize a particular subgroup of patients with the DG-HCM phenotype. Group of patients with any other anatomic variant of cardiac hypertrophy (non-DG-HCM, 136 patients) we used as a group of comparison when possible.

### Clinical and instrumental evaluation

This study was performed in accordance with the Helsinki declaration and the ethics committee of Petrovsky National Research Center of Surgery. Data obtained from each individual in the study included personal and familial medical history, general examination, 12-lead resting ECG, 24-hour ECG Holter monitoring, transthoracic and transesophageal (intra-operative) EchoCG, cardiac computer tomography (CT), cardiac magnetic resonance imaging (MRI) with gadolinium enhancement.

Transthoracic echocardiography (TTE) was made using a Vivid-7 Dimension with a multi-frequency probe 2,5/4,7 MHz, with synchronization of an electrocardiogram. Typical TTE included: M-mode, B-mode, PW, and CW Doppler, color flow Doppler and Doppler tissue imaging (DTI). TTE parameters were recorded at rest and also during functional tests: Valsalva maneuver and exercise test with minimal physical activity (15-20 squats). The TTE was made before the surgical treatment and 7-14 days after surgery. Additionally, we determine systolic flow in the left ventricular (LV) outflow tract (LVOT) and flow velocity pattern in the cavity of LV.

We determined the parameters of LV diastolic, systolic, and global functions at rest and during functional tests. We have made a qualitative and quantitative evaluation of the anatomy and histopathology of the mitral valve (MV), changes in the structure and motion of the valves, its hemodynamic contribution to the intracardiac blood flows, the severity of changes in diastolic trans-mitral flow, and systolic flow of mitral regurgitation. For calculations, images were used to clearly visualize the contours and tip of the heart, atrioventricular valves, interventricular septum, and endocardial borders.

The risk of SCD was evaluated using the “HCM-Risk SCD Calculator” (9). Histological and immunohistochemical assays were performed on the myocardial tissue specimens from all of the resected structures: IVS, papillary muscles, and mitral valves leaflets (if prosthetics).

Initial perioperative data were obtained from the patients’ hospital stay. The follow-up data were collected from patients during annual re-evaluations (when available).

### Genetic analysis

Genetic screening of the ten genes encoding sarcomeric proteins (*MYBPC3, TAZ, TPM1, LDB3, MYL2, ACTC1, MYL3, MYH7, TNNI3*, and *TNNT2*) was performed by IonTorrent PGM® (ThermoFisher Scientific, USA), followed by additional Sanger sequencing of the low-covered regions to confirm all detected variants. Mutational testing in additional genes, causing some certain genetic syndromes (*LAMP2, XGAL, PRKAG2, PTPN11, SOS1*, and *BRAF*) was performed by bidirectional Sanger sequencing.

### Statistical analysis

The results were analyzed using the following statistical methods: 1) analysis of the type of distribution of quantitative data using the Shapiro-Wilk test; 2) determination of sample parameters (mean, median, range, standard deviation, minimum, maximum); 3) comparison of two unrelated groups according to the Mann-Whitney test. Calculations were made using the STATISTICA 12.0 application package. The critical value of the level of statistical significance when testing the null hypothesis was taken equal to 0.05.

## Results

Over the course of 7 years, we evaluated and performed open-heart surgery for 170 patients with entering diagnosis of obstructive HCM.

In our daily practice, we distinguished a particular phenotype of cardiac hypertrophy characterized by diffuse hypertrophy of the left ventricular (LV) free walls, whole interventricular septum, and papillary muscles, with their displacement towards the LV apex (Fig.1). Thirty-four out of 170 (20%) had this particular anatomical The gender ratio in DG-HCM was not different from the other 136 HCM patients (M:F ratio was 18:16 vs. 66:70, the difference was not statistically significant). Patients with DG-HCM (median age 53 years, interquartile range 37-59 years, age_min_ 15 years, age_max_ 66 years) were younger than patients with a non-diffuse form of oHCM (median age 58 years, interquartile range 48-64 years, age_min_ 25 years, age_max_ 77 years), (p=0.0011).

**Figure 1.**
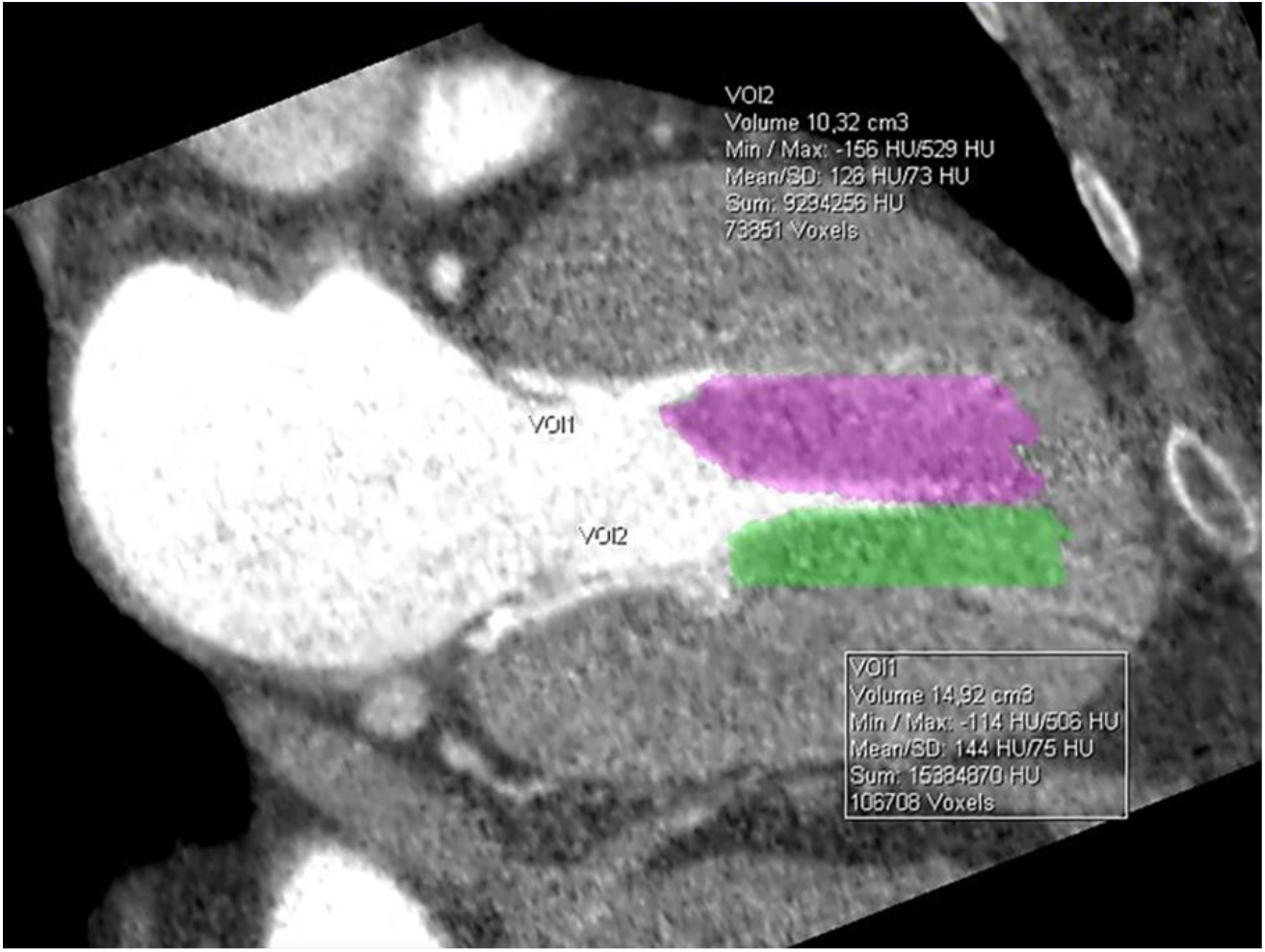
Fragment of the multi-sliced CT. Prominent diffuse hypertrophy of the interventricular septum with expansion to the posterior and lateral walls of the left ventricle. Pronounced hypertrophy and displacement of the bases of the papillary muscles to the apex of the left ventricle. The total volume of the papillary muscles exceeds 25 sm^3^.

Prior to surgery: PGr 71.4±6.7 mmHg, diastolic IVS 25.7±6.2 mm, myocardial mass 309±46 g, and severe mitral regurgitation (3-4 grade) was found in all cases. By NYHA classification, all patients were in the III-IV functional class, and the indications for surgery were considered urgent. Four patients had infection endocarditis in medical history with following mild aortic stenosis (PGr<40 mm Hg). Two patients had subaortic fibrotic membranes. One patient underwent a coronary artery bypass graft (CABG) procedure. Left ventricular noncompaction was revealed in 1 patient.

High five-year risk of SCD (>6%) [9] and extensive myocardial fibrosis (>15% of the septum or free walls) were found in 18 patients with DG-HCM. It was significantly higher than in patients with other anatomic variants (52.9% vs. 9.6%, Fisher exact p, one-tailed <0,0001). With the goal of primary SCD prevention, ICDs were implanted in all 18 patients with a high five-year risk of SCD (9) before or right after open-heart surgery. The follow-up period lasted from 12 to 60 months. Appropriate ICD shocks were registered in 5 out of 18 patients with an implanted devices (28%) during the follow-up period.

Heart transplantation instead of septal myectomy was recommended for three patients because of severe fibrosis, diastolic dysfunction, and progressive heart failure. Thirty-one out of 34 DG-HCM patients underwent reconstructive surgical treatment. An experienced multi-disciplinary team regularly evaluated 29 patients during this period.

Long-term results (up to 5 years) based on annual in-hospital multi-functional examinations were available for 29 out of 31 operated patients.

All reconstructive surgical interventions were performed through a median sternotomy using standard cardiopulmonary bypass (CPB) techniques and hypothermic cardioplegia. Transaortic extended septal myectomy down to the base of the anterior papillary muscle was executed after cardioplegic arrest how it was described [10, 11]. The universal chordal sparing valve replacement with preservation of annulo-papillary contacts in the anterior and posterior annulus semi-circus was performed on 24 patients [11], and submitral plastic reconstruction (parietal resection disengagement of papillary muscles) with mitral valve preservation in 7 patients.

Visual assessment of the left ventricular cavity was performed with excision of the excessive papillary muscles or major apical trabeculae (if present). The volume of the papillary muscles with preserved chordae was decreased via longitudinal parietal resection. In all cases of prosthetics, bicuspid prostheses (“ATS” or “Carbomedics”) were implanted. For six patients (19%) with a significant increase in the size of the left atrium (> 5 cm), volume-reducing plasty of the left atrial posterior wall was performed [11]. There was no complication related to the intra-ventricular stage of reconstructive surgery during or after the operation.

Additional surgical manipulations were performed for seven patients (23%), including coronary artery bypass graft surgery (1 case), subaortic membrane resection (2 cases), and aortic valve replacement due to severe calcinosis (4 cases). In 1 year after surgery, one of the patients who was diagnosed with LEOPARD syndrome (carrier of the p.Thr468Met heterozygous mutation in the *PTPN11* gene) died suddenly out of the hospital. The cause of death was recurrent ventricular tachycardia (electrical storm) despite multiple appropriate shocks from the ICD.

A dramatic decrease in the intraventricular pressure gradient was observed intraoperatively. This trend continued during the follow-up period. Mean intraventricular PGr was 71±6.7 mmHg before surgery versus 4.75±1.3 mmHg during the follow-up period. The mean interventricular septal thickness before surgery was 25.7±6.2 mm versus 15±2.2 mm during the last reevaluation. These differences are statistically significant and therefore support the positive clinical results of the performed reconstructions.

The severe mitral regurgitation (grade 3-4) was successfully corrected with extended submitral reconstruction or after the prosthesis. Left atrial volume was decreased as a result of surgical plastic remodeling in 6 patients and correction of the mitral insufficiency. All patients showed significant clinical improvement and remained in the I-II functional class (by NYHA classification) during the entire follow-up period: shortness of breath at rest and moderate exercises disappeared completely; no dizziness and or syncope were registered during all period of follow-up; medication was limited by beta-blockers and anticoagulants for patients with biological prosthetics. The normalization of cardiac function was also confirmed by the decrease in the BNP level to reference values ranges after surgery (772.7±438 pg/mL before and <60 pg/mL after surgery).

The delayed gadolinium enhancement observed during cardiac MRI with a gadolinium-based contrast medium corresponding to diffuse myocardial fibrosis was found in all patients.

Histopathological examinations had revealed severe fibrosis, cardiomyocytes hypertrophy and disarray, intra-cellular hyperplasia in the samples taken both from intraventricular septum and papillary muscles (Fig.2). All mitral valve leaflets had unusual elongated anatomy with leaflet cooptation zone displacement towards the left ventricular chamber. Histological features consistent with leaflets tissue dysplasia and myxomatous degeneration were found in all specimens.

**Figure 2.**
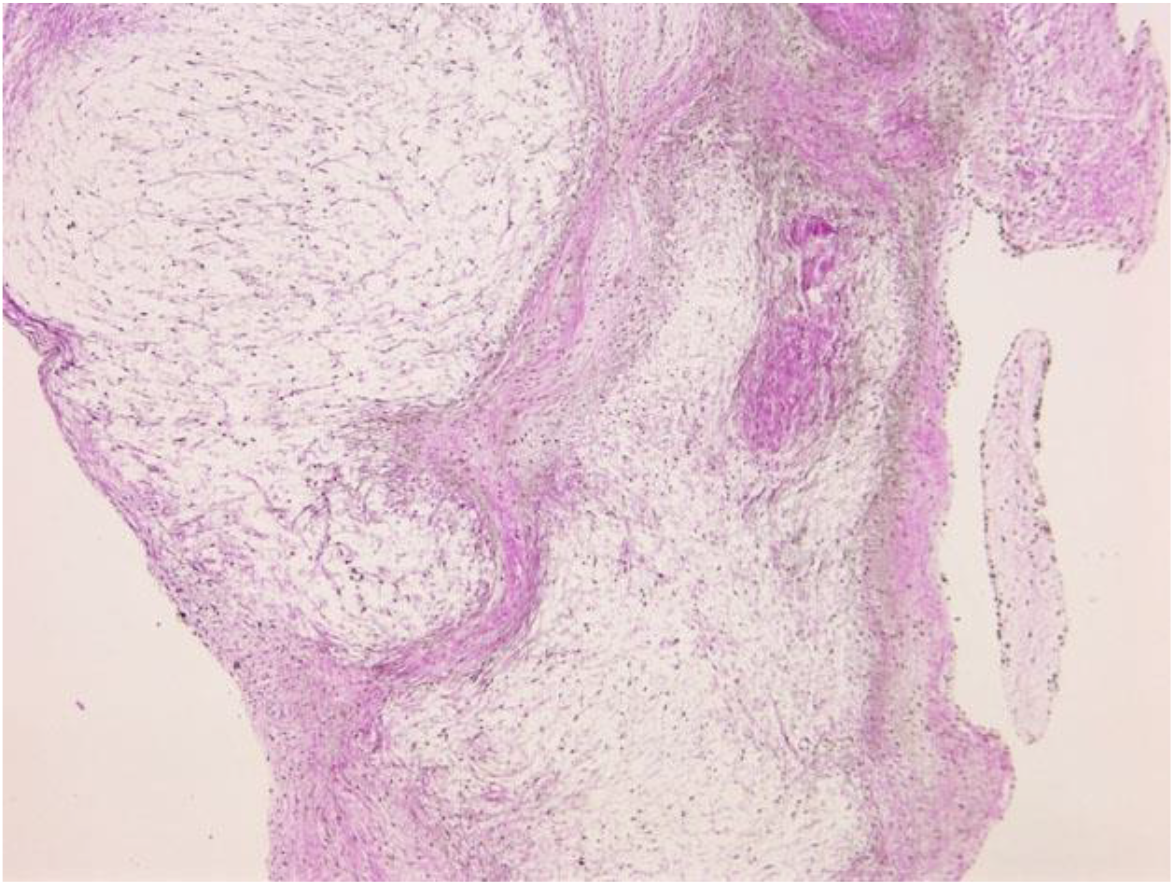
The papillary muscle of diffuse generalized HCM: interstitial fibrosis (van Giesons stain, х100).

The spectrum of genetic causes of LV hypertrophy in the primary cohort of 34 patients with diffuse generalized HCM was analyzed. Unequivocal familial anamnesis of the cardiomyopathy was found in 12 families with DG-HCM (35%). SCD in families was reported by 2 probands without reliable data of cardiac hypertrophy, 12 probands (35%) were self-reported as sporadic, and 8 probands had no relevant information about family history.

Non-sarcomeric inherited disorders associated with LV enlargement (lysosomal storage diseases, RASopathies, and neuromuscular disorders) were found in 12 patients (Fig. 3). None “non-sarcomeric” inherited syndrome was suspected/confirmed in patients with the non-DG-HCM phenotype (12 out 34 pts vs. 0 out 136, Fisher exact p, one-tailed <0,0001).

**Figure 3.**
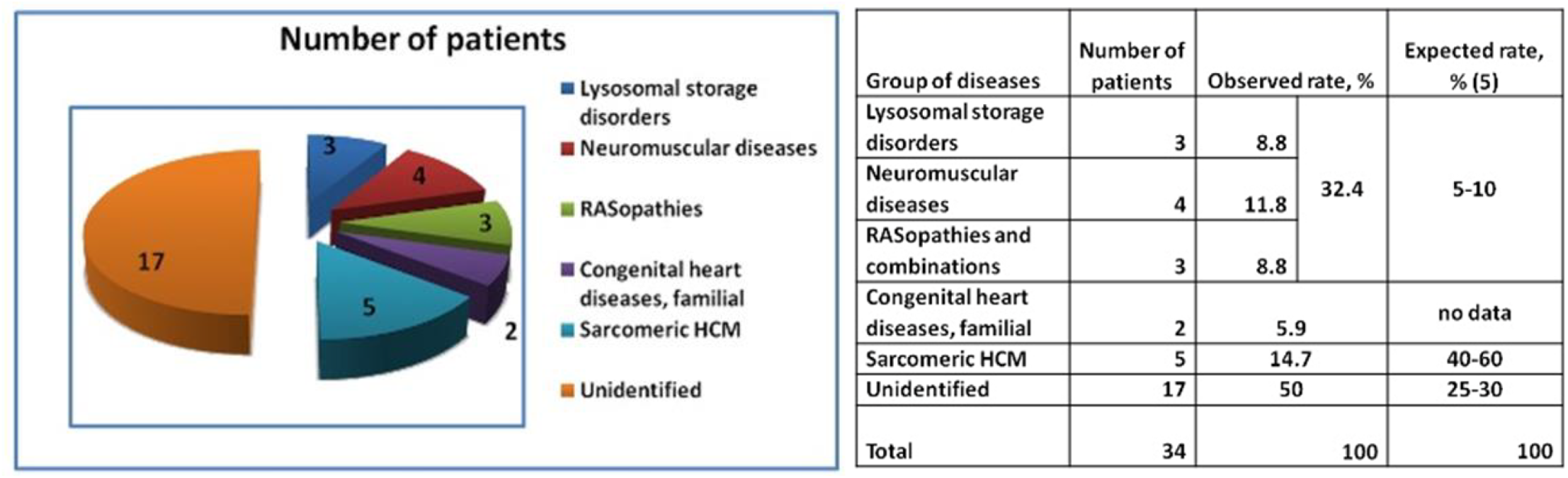
Distribution of the inherited disorders associated with LV found in the group of patients with DG-HCM. The number of probands is shown in the diagram.

One patient out of 34 was diagnosed with left ventricular noncompaction in addition to the prominent cardiac hypertrophy. Orthotopic heart transplantation as an optimal surgical option was recommended for 3 of the patients diagnosed with lysosomal Danon disease.

Mutations in the “classic” genes encoding sarcomeric proteins were only found in 5 out of 34 patients (15%). The spectrum of causative genetic variants is shown in Table 1. Overall, ten rare genetic variants were detected in nine unrelated patients, and pathogenicity was evaluated based on ACMG 2015 criteria [12]. Only five variants (2 in the *MYH7* gene, and 3 in the *MyBPC3* gene) were classified as likely pathogenic or pathogenic (IV-V classes). One patient carried two heterozygous causative variants (p.R502Q in the *MyBPC3* gene, and p.N1327K in the *MYH7* gene). Five genetic variants were classified as VUS (Class III).

**Table 1.** Genetic variants identified in the genes encoding sarcomeric proteins.

| Gene | Substitution (DNA) | Effect (protein) | rs (dbSNP) | Class (ACMG/AMP2015) | References | Criteria of pathogenicity |
| --- | --- | --- | --- | --- | --- | --- |
| *TPM1* | c.775A>G | p.K259Q | rs144045691 | III | CV | VUS: PP3, BS1 |
| *LDB3* | c.608C>T | p.S203L | rs201538257 | III | (16) | VUS: PM2, PP3 |
| *MyBPC3* | c.3197C>G | p.Pro1066Arg | n/d | III | (17) | VUS: PM2, PP1, PP3, PP5 |
| *MYH7* | c.3551A>G | p.Q1184L | rs546586969 | III | CV | VUS: PM2, PP2, PP3 |
| *MYH7* | c.3981C>A | p.N1327K | rs141764279 | III | CV | VUS: PM2, PP2, PP3 |
| *MYH7* | c.5134C>T | p.R1712W | rs121913650 | IV | CV | LP: PM2, PM5, PP2, PP3, PP5, |
| *MYH7* | c.1183C>A | p.L395M | rs1324850187 | IV | CV | LP: PM1, PM2, PP2, PP3 |
| *MyBPC3* | c.3697C>T | p.Q1233* | rs397516037 | IV/V | CV | LP/P: PVS1, PM2, PP5 |
| *MyBPC3* | c.1505G>A | p.R502Q | rs397515907 | V | CV | LP: PM2, PM5, PP1, PP3, PP5 |
| *MyBPC3* | c.1227-13G>A | Affecting splicing | rs397515893 | IV | (18), CV | LP: PS3, PM2, PM4, PP1, PP3, PP5 |

ACMG/AMP - American College of Medical Genetics and Genomics and the Association for Molecular Pathology; CV – references are available in ClinVar database; LP – likely pathogenic; n/d – no data; P – pathogenic; VUS – variant of unknown significance

## Discussion

Current research by several groups is shedding light on the prevalence, genetic etiology, pathogenesis, anatomical subtypes, and clinical polymorphism of HCM. Increasing knowledge of the anatomical subtypes of HCM is leading to the development of new surgical techniques and a better understanding of which patients would benefit from them, thus improving the long-term prognosis for many affected individuals [13]. Decision making in HCM involves accurate diagnosis, the timing of intervention, SCD risk assessment, and further selection of the best interventional technique for every patient. It was shown that tight differentiation of intra-cardiac anatomic phenotypes in oHCM may be significant for the clinical decision making [14]. Four subtypes of septum morphology in hypertrophic heart were distinguished by authors from Mayo’s Clinic based on the echocardiography-contour contour of the interventricular septum in 2006 [7]. Nowadays, cardiac CT and MRI become a part of routine evaluation, and can provide new insights into complex intra-cardiac morphology, especially papillary muscles, additional trabeculae, and mitral valve dysfunction.

Summarizing anatomical, histological, and functional features of diffuse generalized HCM, we particularly mark the following traits:

- Septal (predominantly middle portion) and apical hypertrophy of the LV;
- An excessive number of papillary muscles, their prominent hypertrophy, and dislocation to the apex of the LV;
- Significant decrease in left ventricular diastolic volume with marked diastolic dysfunction;
- Elongated mitral leaflets and cooptation zone migration towards the left ventricular apex, grade 3+ or 4+ mitral insufficiency, and connective tissue dysplasia of the mitral valve;
- Left atrial enlargement with a high risk of developing atrial fibrillation;
- Pulmonary hypertension;
- Pronounced myocardial fibrosis with a high risk of the SCD due to ventricular arrhythmias;
- High risk of progression toward HF.

Genetic testing revealed a lower than expected rate of mutations in the genes encoding sarcomeric proteins (15% of patients) as compared to the overall prevalence of all of the mutations (70-75%) in the HCM cohort [5], and as compared to the expected mutation rate in genes encoding sarcomeric proteins (40-60%) [5]. It is important to note that mutation rates in 2014 ESC guidelines were published before the clinical implication of ACMG criteria. Therefore, a high expected rate (40-60%) of mutations in genes encoding sarcomeric proteins could be partly attributable to the misinterpretation of genetic variants. This explanation includes the differences in rates of unidentified cases (25-30% in 2014 ESC guidelines versus 44.1% in our study). Some genetic variants were classified as variants of uncertain clinical significance due to a lack of reliable family history or functional studies.

Non-sarcomeric causes of clinically significant myocardial hypertrophy (inherited syndromes, neuromuscular disorders, and lysosomal storage diseases) were found in one-third of index DG-HCM cases, an amount also much higher than expected based on guidelines [5]. It is possible that patients with inherited syndromes or lysosomal storage diseases tend to develop more severe clinical symptoms and, therefore, have a higher probability of being referred to a cardiosurgery department. But it is important to note that three patients with lysosomal disorders (Danon disease) of this group who had the most severe diffuse cardiac hypertrophy did not undergo myectomy, and were referred to the heart transplantation waiting list.

Overall, the described phenotype could be found in patients with hypertrophy of various etiologies. The genetic background of each individual likely plays an important role in the anatomical subtype of LV hypertrophy and influences the natural course of cardiac hypertrophy and the long-term prognosis.

The results of the histopathological examinations of the myocardium samples and mitral valve leaflets are of particular interest. They revealed extensive connective tissue dysplasia in 100% of the cases. The co-existence of DG-HCM and connective tissue dysplasia resulting from single genetic mutations might highlights the common genetic and molecular mechanisms underlying normal cardiac and valvular development [15].

The presence of cardiomyocyte hypertrophy and intracellular hyperplasia may help to explain the findings of papillary muscle enlargement and apical myocardial thickening, which cannot be explained by the functional overload. This genetically determined cardiomyocytes hypertrophy plays a significant role in the HCM pathogenesis.

The high level of fibrosis in patients with diffuse generalized HCM has been well documented. Further research is needed to investigate the cross-correlation between the grade of intramyocardial fibrosis, the late gadolinium enhancement in cardiac MR, and long-term prognosis.

The findings in this study emphasize the need for further specification of SCD risk factors, including family history, the role of particular genetic mutations, the degree of hypertrophy/hyperplasia, and the severity of fibrosis. Detailed risk stratification is crucial for the prevention of SCD and to outline the indications for ICD.

The surgical reconstructive technique requires transaortic access for extended septal myectomy and atrial access to perform sub-mitral papillary muscles reconstruction. This technique is effective and can alleviate symptoms of HCM and heart failure. However, it is more effective when performed before the left atrial enlargement and atrial fibrillation.

Although surgical intervention (septal myectomy or alcohol septal ablation) remains an effective treatment for symptomatic HCM patients, an important pharmacological alternative has now become available with the new class of drugs such as cardiac myosin inhibitors. Mavacamten (formerly MYK-461), the first in-class drug approved for clinical use, selectively reduces excessive myocardial contractility, thereby decreasing the left ventricular outflow tract pressure gradient and improving symptoms, exercise tolerance, and quality of life in many patients. Data from the EXPLORER-HCM and VALOR-HCM trials indicate that mavacamten can enable a substantial proportion of patients previously considered candidates for septal reduction therapy to defer or to avoid surgery for significant period of time [19, 20]. Similar agents, such as aficamten, have shown comparable efficacy. However, treatment is not universally tolerated: some patients experience adverse effects, including reductions in left ventricular ejection fraction or other intolerance, necessitating dose adjustment or discontinuation. In such cases, surgical intervention may ultimately prove to be the more appropriate and definitive option. Thus, while cardiac myosin inhibitors expand the therapeutic arsenal for obstructive hypertrophic cardiomyopathy, individualized assessment remains essential.

## Conclusion

The anatomic variant of HCM may correlate with the genetic cause of cardiac hypertrophy and long-term prognosis. Particular diffuse generalized phenotype accounts for 20% of HCM patients with LVOT obstruction who met criteria for the open-heart surgery. Reconstructive extended myectomy with parietal resection of the papillary muscles provides several long-term benefits for patients with DG-HCM reducing the risk of SCD and restoring the functional class close to normal. An excessive level of fibrosis may explain the high risk of ventricular fibrillation and indicate the need for early reconstructive surgery as primary prevention of SCD.

Biomarkers such as genetic mutations and fibrosis level look very promising in the improvement of the SCD risk assessment and long-term prognosis for HF development. Multisystem disorders (neuromuscular disorders, lysosomal storage diseases, RASopathies, etc.) were enriched up to 30% in this group. So special attention should be paid to do not overlook a phenocopy of HCM in this group of patients. It can be of special importance because many of these diseases have additional gene-specific treatment options and/or unique gene-specific recommendations for the long-term follow-up.

## Limitations

This is a relatively small study in the number of patients with diffused generalized HCM: this does not allow us to establish strong phenotype-genotype correlations. The need for emergency surgery (implantation of ICD and open intracardiac correction) in this challenging subgroup of patients precludes the possibility of evaluating a control comparison group requiring only medication.

## Highlights

- Hypertrophic cardiomyopathy is a clinically and genetically complex pathology with variations in clinical manifestation, prognosis, and treatment options. Intra-cardiac anatomy also varies, and several common phenotypes can be characterized based on instrumental data.
- In this observational study, we describe a particular phenotype, diffuse generalized form of HCM, which is distinct from isolated basal hypertrophy of interventricular septum leading to the LVOT obstruction.
- It often requires ICD implantation and special surgical technique for normalization of intracardiac hemodynamics: increasing the LV cavity by extended septal myectomy, parietal resection of the enlarged papillary muscles and their disengagement, and mitral regurgitation correction.
- The spectrum of genetic findings in this group was enriched up to 30% by multisystem disorders (neuromuscular disorders, lysosomal storage diseases, RASopathies, etc.).
- It can be of special importance because many of these diseases have additional gene-specific options for treatment or special gene-specific recommendations for the long-term follow-up; therefore, special attention should be paid to do not overlook a phenocopies of HCM in this group of patients.

## Data Availability Statement

All data produced in the present study are available upon reasonable request to the authors

## Abbreviations and Acronyms

CPB: cardiopulmonary bypass
ECG: electrocardiography
ICD: implantable cardioverter-defibrillator
IVS: interventricular septum
HCM: hypertrophic cardiomyopathy
LEOPARD acronym: (L) stands for (L)entigines; (E)lectrocardiographic rhythm and/or conduction defects; (O)cularhypertelorism; (P)ulmonary stenosis; (A)bnormalities of the genitals; (R)etarded growth; and (D)eafness or hearing loss.
LV: left ventricle
MRI: magnetic resonance imaging
NYHA: New York Heart Association
PGr: intraventricular pressure gradient
Pts: patients
SCD: sudden cardiac death
SD: standard deviation
VT: ventricular tachycardia

## References

1. Semsarian C, Ingles J, Maron MS, Maron BJ. New Perspectives on the Prevalence of Hypertrophic Cardiomyopathy. Journal of the American College of Cardiology. 2015 Mar;65(12):1249–54.

2. Spirito P, Autore C, Formisano F, Assenza GE, Biagini E, Haas TS, et al. Risk of Sudden Death and Outcome in Patients With Hypertrophic Cardiomyopathy With Benign Presentation and Without Risk Factors. The American Journal of Cardiology. 2014 May;113(9):1550–5.

3. Maron BJ, Braunwald E. Evolution of Hypertrophic Cardiomyopathy to a Contemporary Treatable Disease. Circulation. 2012 Sep 25;126(13):1640–4.

4. Maron BJ, Maron MS. A Discussion of Contemporary Nomenclature, Diagnosis, Imaging, and Management of Patients With Hypertrophic Cardiomyopathy. The American Journal of Cardiology. 2016 Dec;118(12):1897–907.

5. 2014 ESC Guidelines on diagnosis and management of hypertrophic cardiomyopathy: The Task Force for the Diagnosis and Management of Hypertrophic Cardiomyopathy of the European Society of Cardiology (ESC). European Heart Journal. 2014 Oct 14;35(39):2733–79.

6. Schaff HV, Dearani JA, Ommen SR, Sorajja P, Nishimura RA. Expanding the indications for septal myectomy in patients with hypertrophic cardiomyopathy: Results of operation in patients with latent obstruction. The Journal of Thoracic and Cardiovascular Surgery. 2012 Feb;143(2):303–9.

7. Binder J, Ommen SR, Gersh BJ, Van Driest SL, Tajik AJ, Nishimura RA, et al. Echocardiography-Guided Genetic Testing in Hypertrophic Cardiomyopathy: Septal Morphological Features Predict the Presence of Myofilament Mutations. Mayo Clinic Proceedings. 2006 Apr;81(4):459–67.

8. Dzemeshkevich SL, Frolova YuV, Kim SYu, Fedorov DN, Zaklyazminskaya EV, Fedulova SV, et al. Anatomic and morphological signs of a diffuse-generalized hypertrophic cardiomyopathy. Russ J Cardiol. 2015 Jan 1;(5):58–63.

9. HCM Risk-SCD Calculator [Internet]. HCM Risk-SCD Calculator. Available from: https://doc2do.com/hcm/webHCM.html

10. Schaff HV, Said SM. Transaortic Extended Septal Myectomy for Hypertrophic Cardiomyopathy. Operative Techniques in Thoracic and Cardiovascular Surgery. 2012;17(4):238–50.

11. Dzemeshkevich S, Korolev S, Frolova J, Skridlevskaya E, Margolina A, Podlesskich Y, et al. Isolated replacement of the mitral leaflets and “Mercedes”-plastics of the giant left atrium: surgery for patients with left ventricle dysfunction and left atrium enlargement. J Cardiovasc Surg (Torino). 2001 Aug;42(4):505–8.

12. Richards S, Aziz N, Bale S, Bick D, Das S, Gastier-Foster J, Grody WW, Hegde M, Lyon E, Spector E, Voelkerding K, Rehm HL; ACMG Laboratory Quality Assurance Committee. Standards and guidelines for the interpretation of sequence variants: a joint consensus recommendation of the American College of Medical Genetics and Genomics and the Association for Molecular Pathology. Genet Med. 2015 May;17(5):405–24. doi: 10.1038/gim.2015.30. Epub 2015 Mar 5. PMID: 25741868; PMCID: PMC4544753.

13. Wang S, Cui H, Yu Q, Chen H, Zhu C, Wang J, et al. Excision of anomalous muscle bundles as an important addition to extended septal myectomy for treatment of left ventricular outflow tract obstruction. The Journal of Thoracic and Cardiovascular Surgery. 2016 Aug;152(2):461–8.

14. Quintana E, Sabate-Rotes A, Maleszewski JJ, Ommen SR, Nishimura RA, Dearani JA, et al. Septal myectomy after failed alcohol ablation: Does previous percutaneous intervention compromise outcomes of myectomy? The Journal of Thoracic and Cardiovascular Surgery. 2015 Jul;150(1):159–167.e1.

15. Rumyantseva V, Zaklyazminskaya E. Clinical and genetic variety of hereditary connective tissue dysplasia. Clin Experiment Surg Petrovsky J. 2015;(2):5–17.

16. Atlas of Genetic Variation [Internet]. Available from: https://www.cardiodb.org/acgv/acgv_variant.php?id=63016

17. Niyazova SS, Chakova NN, Komissarova, SM, Sasinovich MA. Mutation spectrum in sarcomeric protein genes and their phenotypic features in Belarusian patients with hypertrophic cardiomyopathy. Medical Genetics. 2019;6(204):21–12.

18. Jääskeläinen P, Kuusisto J, Miettinen R, Kärkkäinen P, Kärkkäinen S, Heikkinen S, et al. Mutations in the cardiac myosin-binding protein C gene are the predominant cause of familial hypertrophic cardiomyopathy in eastern Finland. J Mol Med. 2002 Jul;80(7):412–22.

19. Olivotto I, Oreziak A, Barriales-Villa R, et al. Mavacamten for treatment of symptomatic obstructive hypertrophic cardiomyopathy (EXPLORER-HCM): a randomised, double-blind, placebo-controlled, phase 3 trial. Lancet. 2020;396(10253):759–769. doi:10.1016/S0140-6736(20)31792-X.

20. Desai MY, Owens A, Geske JB, et al. Myosin Inhibition in Patients With Obstructive Hypertrophic Cardiomyopathy Referred for Septal Reduction Therapy. J Am Coll Cardiol. 2022;80(2):95–108. doi:10.1016/j.jacc.2022.04.048.

